# High-performing Multi-task Model of Urinary Tract Dilation (UTD) Classification for Neonatal Ultrasound Reports Through Natural Language Processing

**DOI:** 10.1101/2024.01.23.24301680

**Authors:** Yining Hua, Anudeep Mukkamala, Carlos Estrada, Michael Lingzhi Li, Hsin-Hsiao Scott Wang

## Abstract

**Objective:** The urinary tract dilation (UTD) classification system provides objective assessment relevant to hydronephrosis management for children. However, the lack of uniform language regarding UTD in radiology reports leads to significant difficulty in both clinical management and research. We seek to develop a unified multi-task/multi-class model that can effectively extract UTD components and classifications from early postnatal ultrasound (US) reports.

**Methods:** Radiology records from our institution were reviewed to identify infants aged 0-90 days undergoing early ultrasound for antenatal UTD. The report and images were reviewed by the study team to create the ground truth of UTD classification and components (primary outcome). Bio_ClinicalBERT, a variant of the Bidirectional Encoder Representations from Transformers (BERT) model, was used as the embedding layers of the classification model. The model was fine-tuned with 11 linear classification layers. All but the last BERT layer were frozen during the fine-tuning process. The model performance was evaluated with five-fold cross-validation with an 80:20 train-test ratio.

**Results:** 2460 early (0-90 days) US reports were included. The five-fold cross-validated model performance is satisfactory (Weighted F1 > 0.9 for all UTD components). We report the weighted F1 scores, accuracies, and standard deviations for all 11 tasks and their average performance.

**Conclusions:** By applying deep state-of-the-art NLP neural networks, we developed a high-performing, efficient, and scalable solution to extract UTD components from unstructured ultrasound reports using one single multi-task model. This can potentially help standardize and facilitate large-scale computer vision research for pediatric hydronephrosis. Key Words: machine learning, efficiency, ambulatory care, forecasting

## INTRODUCTION

Urinary tract dilation (UTD), defined as hydronephrosis or hydroureter, is a common (1 in 200-300 during prenatal ultrasound evaluation with 0.15-0.67% prevalence in the general population)^1^ and potentially significant clinical finding in children that requires accurate assessment for appropriate clinical management. Radiologic evaluations by ultrasound play a critical role in evaluating UTD. In order to standardize the evaluation of UTD, there are several attempts to provide a more objective and universal grading system for UTD. The most commonly noted ones included the Society of Fetal Urology (SFU)^2^ grading and UTD grading.^3^ SFU grading is simple and easy to use. However, the fact that it does not account for salient details like ureteral dilation, bladder abnormalities, and renal parenchymal evaluations limits its clinical generalizability. On the other hand, the UTD classification system has gained significant popularity with more comprehensive evaluations for both upper and lower tracts.

Accurate identification and classification of UTD components are crucial for effective management and treatment planning. However, the variability in reporting styles and inconsistent terminology used in radiology reports hinder efficient data extraction and analysis. Consequently, there is a need for methods that can reliably and consistently extract UTD-related information from unstructured ultrasound reports and provide standardized and actionable data. To manually correct and categorize the radiology report would require a significant commitment of resources and is unlikely to be feasible as a long-term solution. To address these challenges, we propose a novel approach that leverages advanced natural language processing (NLP) techniques and deep-learning-enabled pre-trained language models to develop a unified multi-task/multi-class model for extracting UTD components and classifications from early postnatal ultrasound reports. Given the success of these techniques in clinical prediction tasks, ^4–9^ we hypothesize that we can develop an NLP model that can effectively and accurately identify UTD classification components and grading from unstructured ultrasound reports.

## MATERIALS AND METHODS

### Data Source and Outcome Definition

The overall study design and workflow is shown in Figure 1. We reviewed ultrasound radiology reports from Boston Children’s Hospital, a freestanding acute care children’s hospital located in Boston, Massachusetts, to identify infants aged 0-90 days who underwent early ultrasound for antenatal urinary tract dilation (UTD) from 2010-2022. The report and images were carefully examined by the study team to create the ground truth for UTD classification and its components, which served as the primary outcome. A total of 2,500 early postnatal ultrasound reports (0-90 days) out of over 14,817 reports were randomly selected for the study cohort. Each report in the study dataset includes information such as the PACS accession number, medical record number (MRN), date of imaging, date of birth, and radiology report texts. We removed cases with invalid MRNs or accession numbers, missing notes, or uncertain annotations. The final study cohort consists of 2460 unique radiology reports.

**Figure 1.**
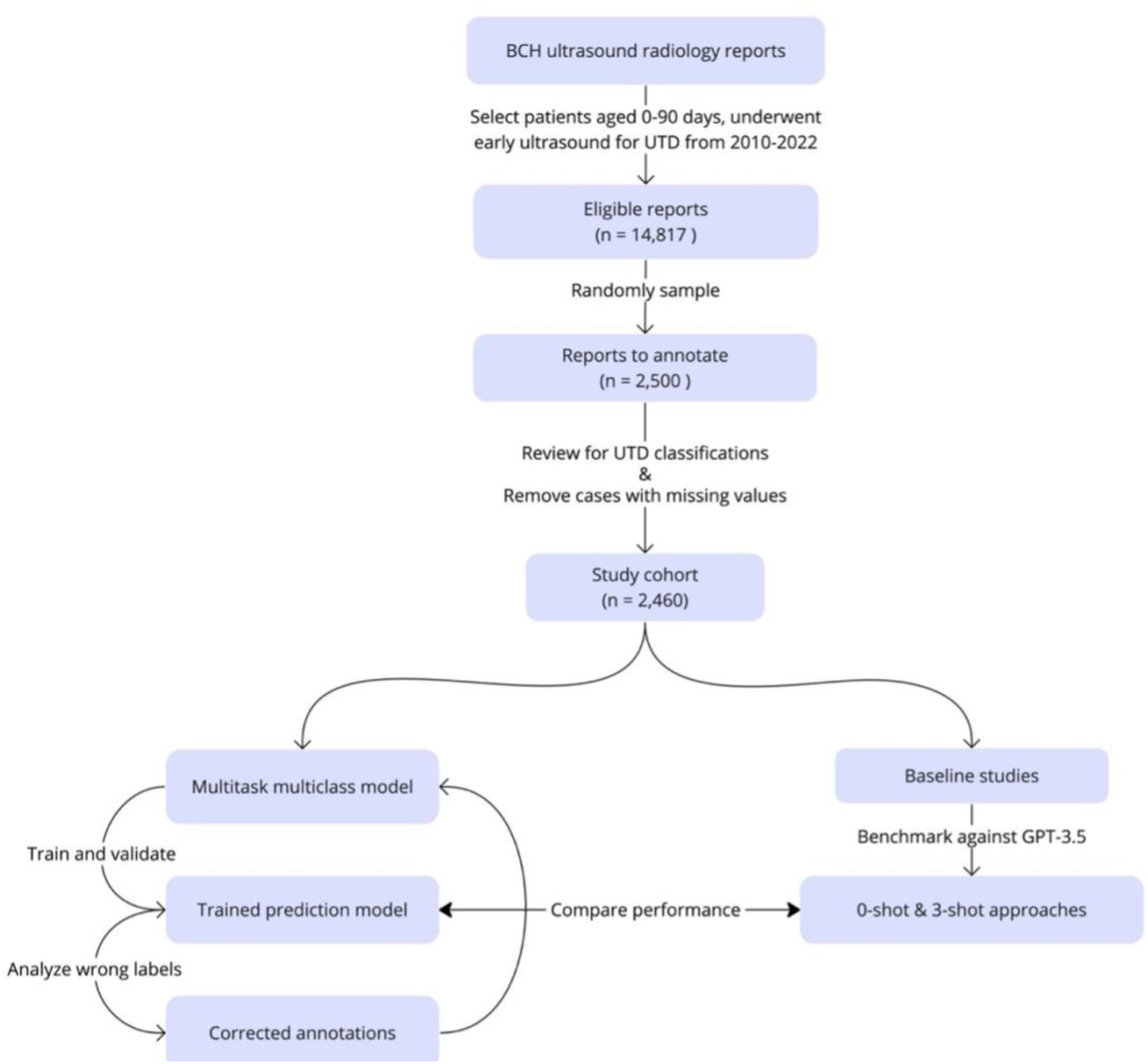
Overall study design and workflow.

We then removed HIPPA-level sensitive information and unrelated details. This involved stripping dates, times, patient IDs, and zip codes, as well as numerical figures with over three digits. We also excluded content data tied to hospital names or places specific to our system.

We defined our outcome (labels) by the UTD classification of 11 tasks for abnormalities including central calyceal dilation (left/right), peripheral calyceal dilation (left/right), abnormal ureter (left/right), abnormal parenchymal thickness (left/right), abnormal parenchymal appearance (left/right), and bladder abnormality. For each task, we defined 3 possible classes: yes, no, or absence (defined as absent kidney or multi-cystic dysplastic kidney). All labels were created by our research team after reviewing the reports as well as the images.

This study has been approved by the Boston Children’s Hospital Institutional Review Board.

### Model Development

The multi-task model consisted of a Bidirectional Encoder Representations from Transformers (BERT)^10^ encoder, followed by task-specific linear classification layers for each of the 11 UTD classification tasks. The decision to adopt BERT was motivated by its outstanding performance across various NLP fields and its relatively compact size compared to other large language models (LLMs). BERT’s contextualized word representations have consistently achieved state-of-the-art results in NLP applications. Bio_ClinicalBERT^11^, a variant of BERT trained on a biomedical corpus that includes radiology notes, was specifically selected, which enables the model to benefit from domain-specific knowledge and improved performance in radiology note comprehension.

These linear layers were connected to the BERT encoder, allowing the model to learn task-specific representations and make predictions. During the fine-tuning process, the BERT layers, except the last one, were frozen to preserve the pre-trained weights and focus on adapting the model to the specific UTD classification tasks.

In our experiments, we employed a stratified sampling technique specifically to address the challenge posed by the imbalanced distribution of our three outcome classes. Stratified sampling ensured that each fold in the 5x5 cross-validation had a representative consistent proportion of each class, thereby improving the model’s ability to generalize across different subsets of the data. WeWe conducted analysis through 5x5 cross-validation, each iteration was initiated with a random seed number to enhance the experiment robustness and reliability. We employed a stratified sampling technique to ensure that each fold of the cross-validation had a consistent proportion of each outcome class, thereby improving the model’s ability to generalize across different subsets of the data. In each iteration, we used an 80:20 ratio for train-test splits. The training phase spanned 25 epochs with a batch size of 12, and the Adam optimizer was configured with a learning rate of 5e-5. Within the training set, we performed a further division into training and validation subsets, adhering to an 80:20 ratio. To monitor the model’s progress and mitigate the risk of overfitting, early stopping was implemented with a patience of 5 epochs.

Finally, we assessed the model’s performance on the out-of-sample test sets using a comprehensive set of evaluation metrics including weighted accuracy, F1 score, precision, recall, the area under the ROC curve (AUC), and its 95% confidence interval (CI) calculated supported by bootstrap resampling on the test sets (n = 1,000).

### Semi-supervised Learning and Error analysis

During the evaluation phase, it came to our attention that a few initial labels were questionable, prompting the adoption of a semi-supervised learning approach to iteratively rectify these labels and enhance the model’s performance in our study. To correct the labels, the model was initially trained on the collected data. Afterwards, instances where the model produced incorrect predictions across five cross-validation iterations were identified for error analysis and relabeling. These relabeled instances, along with the remaining dataset, were then utilized to retrain the model. This iterative process was repeated two times to show the efficiency of this approach, progressively refining the label accuracy, and advancing the overall model performance.

### Experiment Against Existing Large Language Model (LLM)

In our study, we benchmark our algorithm against GPT-3.5, a state-of-the-art LLM developed by OpenAI and commercially available as “ChatGPT.” This model is distinguished for its extensive training data and exceptional capabilities in diverse language-based tasks. For a thorough comparison, we employed GPT-3.5-Turbo, the most advanced version available. We used both “0-shot” and “3-shot” approaches for task execution following previous literature.^12,13^ In the “0-shot” method, we did not provide the model with any task-specific examples, relying solely on its pre-existing training. Conversely, the “3-shot” method included three task-specific examples to facilitate the model’s understanding and performance. These examples were randomly sampled, one from each possible outcome category, to provide a comprehensive task primer. Prompts used in this experiment are provided in Appendix S1 and S2.

### Software and Hardware

An alpha of 0.05 and 95% confidence intervals (CI) were used as criteria for statistical significance. Analyses were performed using Python 3.9 (package Pandas, Numpy, PyTorch) and NVIDIA Titan V with 12GB of GPU memory to accelerate our machine learning computations.

## RESULTS

### Demographics and Cohort Characteristics

Table 1 shows the demographics and year of imaging distribution of our study cohort. The majority of the cohort were males, constituting 67.68% of the total, while females represented 32.32%. The median age of individuals at the time of their ultrasound was 33 years, with an interquartile range spanning from 22 to 62 years. Imaging frequency showed fluctuations over the years, with a notable peak in 2018 at 12.40% of the total scans.

**Table 1.** Demographics and Ultrasound Trends over Years.

|  |  |  |
| --- | --- | --- |
| **Age (median [IQR])** |  |  |
| 33 [22, 62] |  |  |
| **Gender (count, percentage)** |  |  |
| Female | 795 | 32.32% |
| Male | 1665 | 67.68% |
| **Year of Imaging (count, percentage)** |  |  |
| 2011 | 33 | 1.34% |
| 2012 | 189 | 7.68% |
| 2013 | 186 | 7.56% |
| 2014 | 185 | 7.52% |
| 2015 | 147 | 5.98% |
| 2016 | 132 | 5.37% |
| 2017 | 137 | 5.57% |
| 2018 | 305 | 12.40% |
| 2019 | 255 | 10.37% |
| 2020 | 154 | 6.26% |
| 2021 | 178 | 7.24% |
| 2022 | 75 | 3.05% |

Table 2 shows the distribution of labels pertaining to the 11 UTD classification tasks before model-instructed correction. There is a noticeable class imbalance, with the “No” category often overwhelmingly higher than the “Yes” category. Notably, while many tasks present a balance between left and right abnormalities, asymmetry is evident in “central calyceal dilation” with left at 59.35% and right at 31.50%, and “peripheral calyceal dilation” showing left at 52.76% and right at 26.91%.

**Table 2.** Distribution of the Initial Human-Labeled UTD Classification Labels.

| Task | Yes | No | Absent/MDCK |
| --- | --- | --- | --- |
| **left central calyceal dilation** | 1459 (59.35%) | 960 (39.02%) | 41.0 (1.67%) |
| **left parenchymal appearance abnormal** | 196 (7.97%) | 2223 (90.36%) | 41.0 (1.67%) |
| **left parenchymal thickness abnormal** | 175 (7.11%) | 2244 (91.22%) | 41.0 (1.67%) |
| **left peripheral calyceal dilation** | 1298 (52.76%) | 1121 (45.57%) | 41.0 (1.67%) |
| **left ureter abnormal** | 298 (12.11%) | 2121 (86.22%) | 41.0 (1.67%) |
| **right central calyceal dilation** | 775 (31.50%) | 1641 (66.71%) | 44.0 (1.79%) |
| **right parenchymal appearance abnormal** | 170 (6.91%) | 2246 (91.30%) | 44.0 (1.79%) |
| **right parenchymal thickness abnormal** | 147 (5.98%) | 2269 (92.23%) | 44.0 (1.79%) |
| **right peripheral calyceal dilation** | 663 (26.91%) | 1753 (71.30%) | 44.0 (1.79%) |
| **right ureter abnormal** | 221 (8.98%) | 2195 (89.23%) | 44.0 (1.79%) |
| **bladder abnormal** | 61 (2.48%) | 2351 (95.53%) | 48.0 (1.79%) |

### Model Performance

The model exhibited high F1 scores, ranging from 0.90 to 0.98, across the 11 tasks with tight confidence intervals, suggesting a robust and consistent performance (results shown in Table 3). Among the tasks, ‘bladder abnormality’ achieved the highest F1 score of 0.9796 [95% CI: 0.9688, 0.9904] while ‘left peripheral calyceal dilation’ was on the lower end with an F1 score of 0.9012 [95% CI: 0.8908, 0.9116]. The overall average F1 score for the model was 0.9418 [95% CI: 0.9307, 0.9529]. Similarly, the model demonstrated strong precision and recall for all the tasks, further corroborating the effectiveness of our approach. The AUROC scores also indicated excellent discriminative ability, with an average AUROC of 0.9392 [95% CI: 0.9239, 0.9545].

**Table 3.**
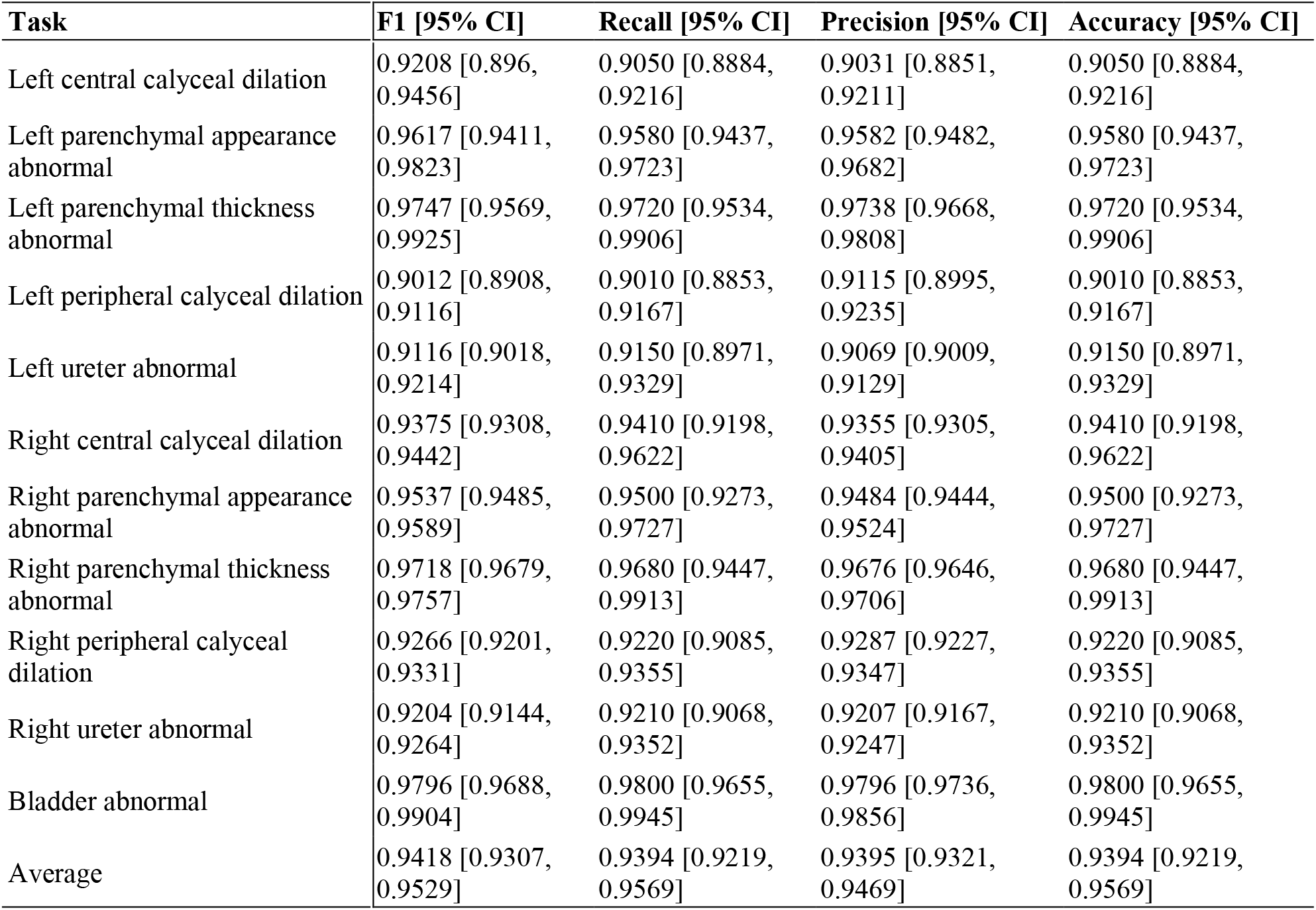
Performance of the Bio_ClincalBERT-based prediction model, round 3.

Figure 2 and Figure 3 present comparative error analyses of human errors and model errors, where ‘Human incorrect’ indicates instances where human labeling was erroneous upon review; ‘Model incorrect’ denotes cases where the model’s predictions were inaccurate; and ‘Human and model incorrect’ represents scenarios where both human labeling and model predictions. We observe that the error analysis from the semi-supervised learning approach reveals distinct error patterns between human and model predictions, with conditions such as ‘left central calyceal dilation’ more commonly mislabeled by humans, while ‘bladder’ errors are more prevalent in model predictions. This suggests that an integrated correction approach leveraging both human and model strengths could improve overall accuracy. Furthermore, the reduction in errors through iterative rounds of label refinement, as reflected by 592 incorrect prediction in round 1 error analysis vs. 529 incorrect predictions in round 2 error analysis, suggests that this method is effective in enhancing the quality of the training data. Appendix S1 and S2 further present the model performance results of rounds 1 and 2 learning and evaluation, from which we observe that training and label correction helped with improving model performance.

**Figure 2.**
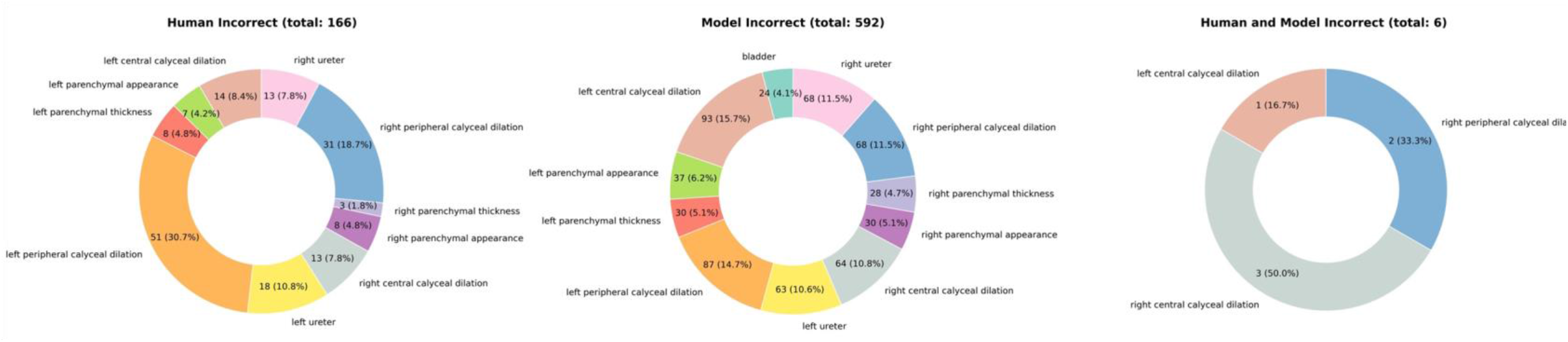
Error Analysis from Round 1 evaluation. ‘Model incorrect’ denotes cases where the model’s predictions were inaccurate; and ‘Human and model incorrect’ represents scenarios where both human labeling and model prediction.

**Figure 3.**
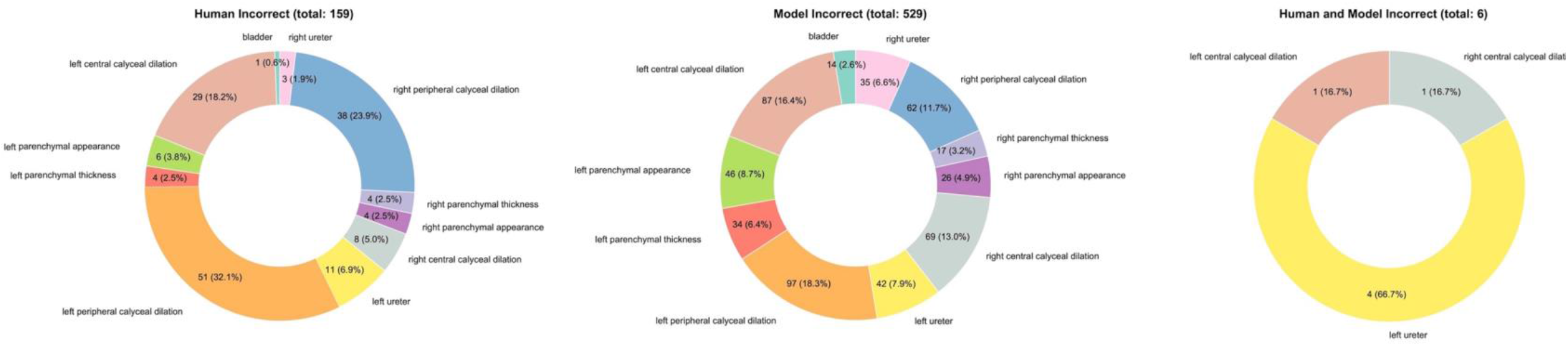
Error Analysis from Round 2 evaluation. ‘Model incorrect’ denotes cases where the model’s predictions were inaccurate; and ‘Human and model incorrect’ represents scenarios where both human labeling and model prediction.

Table 4 shows the baseline results generated by ChatGPT, supported by its best GPT3.5 model at the time (i.e., GPT 3.5-Turbo). In terms of F1, our model outperforms GPT 3.5-Turbo by 20% -30%. Another interesting observation is that providing GPT 3.5 with examples from each of the classes did not help it perform better in the tasks.

**Table 4.**
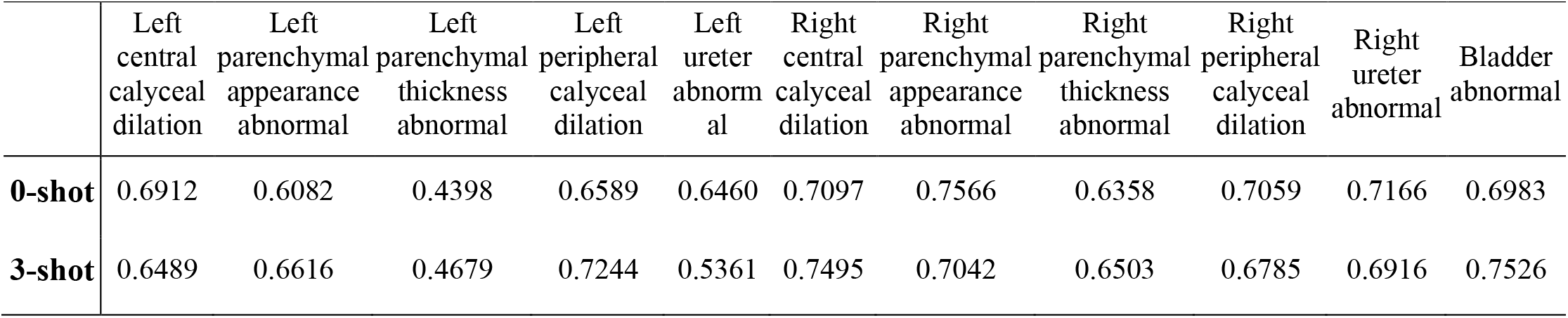
Baseline results using GPT 3.5-Turbo.

|  | Left central calyceal dilation | Left parenchymal appearance abnormal | Left parenchymal thickness abnormal | Left peripheral calyceal dilation | Left ureter abnormal | Right central calyceal dilation | Right parenchymal appearance abnormal | Right parenchymal thickness abnormal | Right peripheral calyceal dilation | Right ureter abnormal | Bladder abnormal |
| --- | --- | --- | --- | --- | --- | --- | --- | --- | --- | --- | --- |
| **0-shot** | 0.6912 | 0.6082 | 0.4398 | 0.6589 | 0.6460 | 0.7097 | 0.7566 | 0.6358 | 0.7059 | 0.7166 | 0.6983 |
| **3-shot** | 0.6489 | 0.6616 | 0.4679 | 0.7244 | 0.5361 | 0.7495 | 0.7042 | 0.6503 | 0.6785 | 0.6916 | 0.7526 |

## DISCUSSIONS

Our proposed approach integrates cutting-edge NLP deep learning algorithms to effectively capture the semantic context and extract meaningful representations from unstructured ultrasound reports. By integrating advanced NLP techniques and semi-supervised learning, we developed a high-performing NLP model to address the challenges associated with UTD classification and component extraction, making notable contributions to the field. The integration of advanced NLP techniques enables our model to effectively comprehend the complex language patterns and nuances present in ultrasound reports. This allows for accurate identification and classification of UTD components, facilitating clinical management and decision-making processes. By automating the extraction of UTD information from ultrasound reports, our approach can provide consistent UTD grading to infants who presented with hydronephrosis. Since UTD grades are well-known to risk categorize hydronephrosis patients, we expect this work would streamline the clinical and research workflow and saves resource.

Another key aspect of our approach is the adoption of semi-supervised learning to rectify incorrect labels. This is crucial in scenarios where labeled data could be prone to errors. By iteratively correcting the labels based on the model’s predictions, we enhance the accuracy and reliability of the training data. This iterative process progressively refines the model’s understanding of UTD components, resulting in improved performance over multiple iterations. Our findings demonstrate the effectiveness of semi-supervised learning in enhancing the model’s capabilities and addressing the challenges inherent in UTD classification.

The implications of our research extend beyond the scope of UTD classification and component extraction. By establishing a standardized and efficient method for extracting UTD information from ultrasound reports, our approach lays the foundation for large-scale machine-learning research on pediatric hydronephrosis. The availability of accurately labeled data enables the development and evaluation of future computer vision algorithms for automated detection, quantification, and analysis of UTD components. This opens up new avenues for research, potentially leading to improved diagnostic accuracy, treatment planning, and patient outcomes.

In addition, the observation that our integrated multi-task human-model substantially outperforms ChatGPT underscores the limitations of relying solely on LLMs. These models, while powerful, are not panaceas; they do not invariably outperform more lightweight models, especially in complex tasks requiring nuanced understanding. The extensive data needed to fine-tune such LLMs, coupled with their intensive computational resource requirements, can be prohibitive. In our case, few-shot learning proved ineffective, possibly introducing noises rather than providing useful information, likely because of the complexity of the study question, involving multiple tasks with a variety of possible classes. This is reflected in our results where our tailored model significantly outperformed GPT 3.5-Turbo by 20% to 30% in F1 scores, indicating that merely providing the GPT model with additional class examples did not enhance its task performance. In real-life expert-domain applications, developing targeted models are likely more efficient than deploying LLMs.

Nonetheless, the results of this study must be interpreted in the context of its limitations. This is a single-institution study based on retrospective review at a tertiary referral center which may limit the generalizability of our findings. Different institutions might have varied patient populations, imaging protocols, or reporting styles, which could influence the outcomes. External validation sets from a different institution or dataset could offer a more rigorous assessment of our model’s broader applicability. Additionally, the process of determining the ground truth for UTD classification, based on the review of reports and images by our research team, introduces another potential source of bias. This subjective approach might lead to errors in labeling, a concern that was substantiated during our evaluation phase when we identified and rectified erroneous annotations. While our model assisted in correcting these annotations, it’s worth noting that if not properly managed, such a process could theoretically reinforce biases, even if we did not observe this trend in our study. Furthermore, the challenge of imbalanced data (i.e., rare labels such as increased parenchymal echogenicity or bladder abnormality) remains a potential concern. Although we employed stratified sampling to mitigate the effects of the imbalanced distribution of our outcome classes, the inherent imbalances in the dataset could still influence the model’s performance. Future studies should consider these limitations and potentially explore multi-institutional datasets and more rigorous validation strategies to enhance the robustness and generalizability of the findings. Lastly, the exclusion of cases with missing values may introduce another layer of bias into our study. By removing these instances, we may inadvertently skew the dataset towards more complete but potentially unrepresentative cases. This could result in a model that is less generalizable to real-world scenarios where missing data are common. For example, cases with missing values might systematically differ from those without, perhaps reflecting more complex or severe conditions that were not fully captured in the dataset. Consequently, the model’s ability to generalize to all types of patients, including those with incomplete records, could be compromised.

## CONCLUSIONS

By applying deep state-of-the-art NLP neural networks, we developed a high-performing, efficient, and scalable solution to extract UTD components from unstructured ultrasound reports using one single multi-task model. This can potentially help standardize and facilitate large-scale computer vision research for pediatric hydronephrosis.

## Data Availability

All data produced in the present study are available upon reasonable request to the authors, pending the necessary institutional reviews and approvals.

## AUTHOR CONTRIBUTIONS

Study design: Yining Hua, Carlos Estrada, Michael Lingzhi Li, Hsin-Hsiao Scott Wang

Data collection and annotation: Anudeep Mukkamala

Experiments: Yining Hua

Analysis: Yining Hua, Michael Lingzhi Li, Hsin-Hsiao Scott Wang

Manuscript drafting: Yining Hua

Manuscript revision and proofreading: All authors.

Yining Hua takes responsibility for the integrity of the work.

## DATA AVAILABILITY STATEMENT

The data and code used for training and evaluation in this study are available upon reasonable request from the corresponding author, pending the necessary institutional reviews and approvals.

## SUPPLEMENTARIES

**S1.**
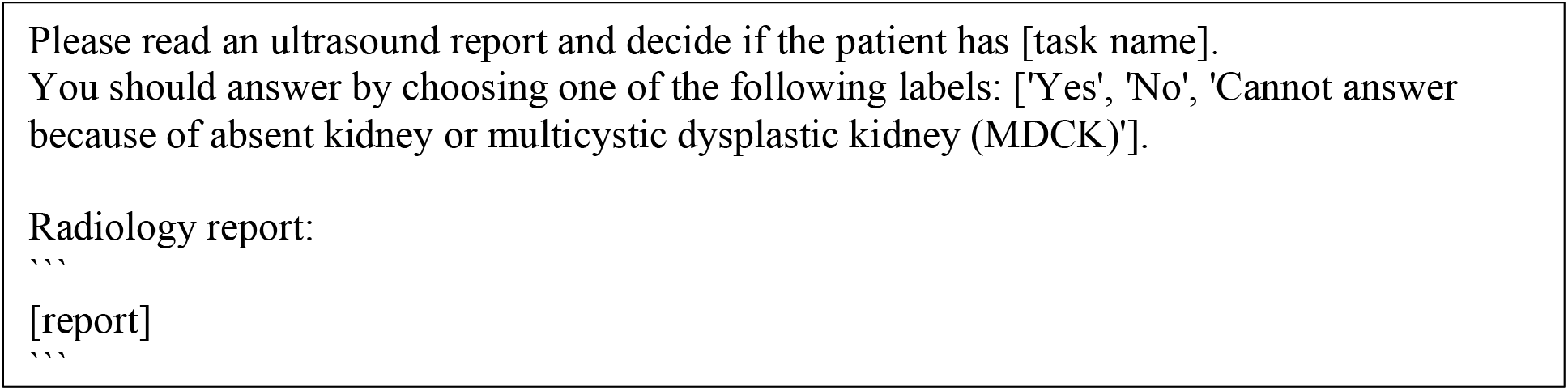
0-shot prompt for GPT-3.5-Turbo. [task name] is a placeholder for tasks, and [report] is a placeholder for actual radiology reports.

**S2.**
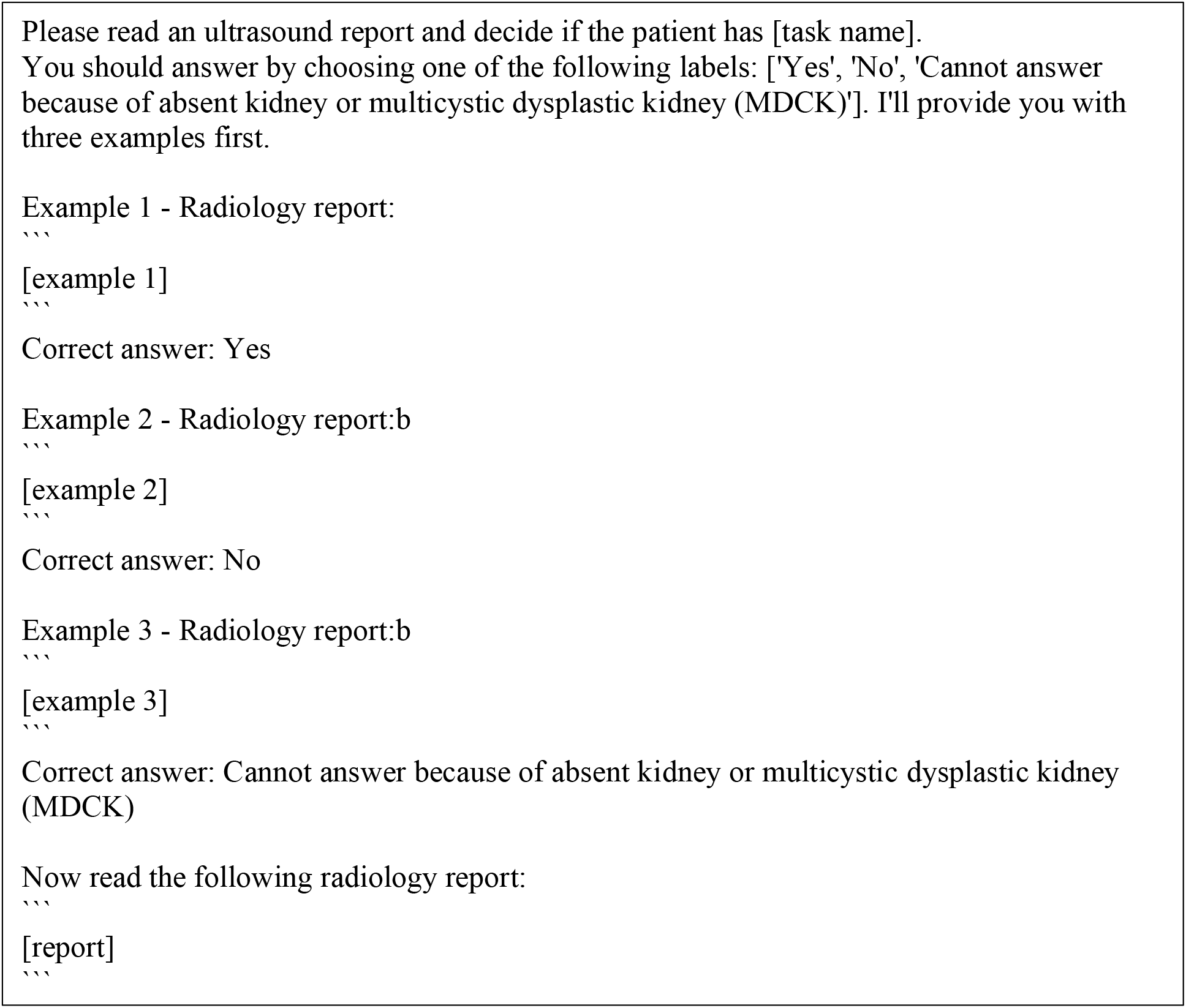
3-shot prompt for GPT-3.5-Turbo. [task name] is a placeholder for tasks, [example n] is a placeholder for example radiology reports matched by the desired task and answer, and [report] is a placeholder for actual radiology reports.

**S3.** Round 1 model performance.

S3. Round 1 model performance
| Task | F1 [95% CI] | Recall [95% CI] | Precision [95% CI] | Accuracy [95% CI] | AUROC [95% CI] |
| --- | --- | --- | --- | --- | --- |
| left central calyceal dilation | 0.9284<br>[0.9032, 0.9536] | 0.9074<br>[0.8902, 0.9246] | 0.8833<br>[0.8651, 0.9015] | 0.9074<br>[0.8902, 0.9246] | 0.9201<br>[0.8948, 0.9454] |
| left parenchymal appearance abnormal | 0.9532<br>[0.9333, 0.9731] | 0.9613<br>[0.9477, 0.9749] | 0.9489<br>[0.9386, 0.9592] | 0.9613<br>[0.9477, 0.9749] | 0.9423<br>[0.9224, 0.9622] |
| left parenchymal thickness abnormal | 0.9645<br>[0.9464, 0.9826] | 0.9715<br>[0.9533, 0.9897] | 0.9610<br>[0.9535, 0.9685] | 0.9715<br>[0.9533, 0.9897] | 0.9518<br>[0.9392, 0.9644] |
| left peripheral calyceal dilation | 0.9060<br>[0.8953, 0.9167] | 0.8841<br>[0.8688, 0.8994] | 0.8771<br>[0.8650, 0.8892] | 0.8841<br>[0.8688, 0.8994] | 0.9026<br>[0.8886, 0.9166] |
| left ureter abnormal | 0.9243<br>[0.9153, 0.9333] | 0.9212<br>[0.9017, 0.9407] | 0.9038<br>[0.8975, 0.9101] | 0.9212<br>[0.9017, 0.9407] | 0.9061<br>[0.8868, 0.9254] |
| right central calyceal dilation | 0.9247<br>[0.9185, 0.9309] | 0.9252<br>[0.9027, 0.9477] | 0.9305<br>[0.9251, 0.9359] | 0.9252<br>[0.9027, 0.9477] | 0.9316<br>[0.9287, 0.9345] |
| right parenchymal appearance abnormal | 0.9502<br>[0.9457, 0.9547] | 0.9541<br>[0.9321, 0.9761] | 0.9478<br>[0.9435, 0.9521] | 0.9541<br>[0.9321, 0.9761] | 0.9465<br>[0.9348, 0.9582] |
| right parenchymal thickness abnormal | 0.9672<br>[0.9628, 0.9716] | 0.9681<br>[0.9452, 0.9910] | 0.9687<br>[0.9655, 0.9719] | 0.9681<br>[0.9452, 0.9910] | 0.9648<br>[0.9564, 0.9732] |
| right peripheral calyceal dilation | 0.9050<br>[0.8982, 0.9118] | 0.9006<br>[0.8868, 0.9144] | 0.9135<br>[0.9074, 0.9196] | 0.9006<br>[0.8868, 0.9144] | 0.9139<br>[0.9083, 0.9195] |
| right ureter abnormal | 0.9316<br>[0.9250, 0.9382] | 0.9275<br>[0.9133, 0.9417] | 0.9278<br>[0.9235, 0.9321] | 0.9275<br>[0.9133, 0.9417] | 0.9302<br>[0.8799, 0.9805] |
| bladder abnormal | 0.9736<br>[0.9615, 0.9857] | 0.9821<br>[0.9653, 0.9989] | 0.9782<br>[0.9719, 0.9845] | 0.9821<br>[0.9653, 0.9989] | 0.9803<br>[0.9749, 0.9857] |
| Average | 0.9390<br>[0.9278, 0.9502] | 0.9366<br>[0.9181, 0.9551] | 0.9310<br>[0.9234, 0.9386] | 0.9366<br>[0.9181, 0.9551] | 0.9355<br>[0.9199, 0.9511] |

**S4.**
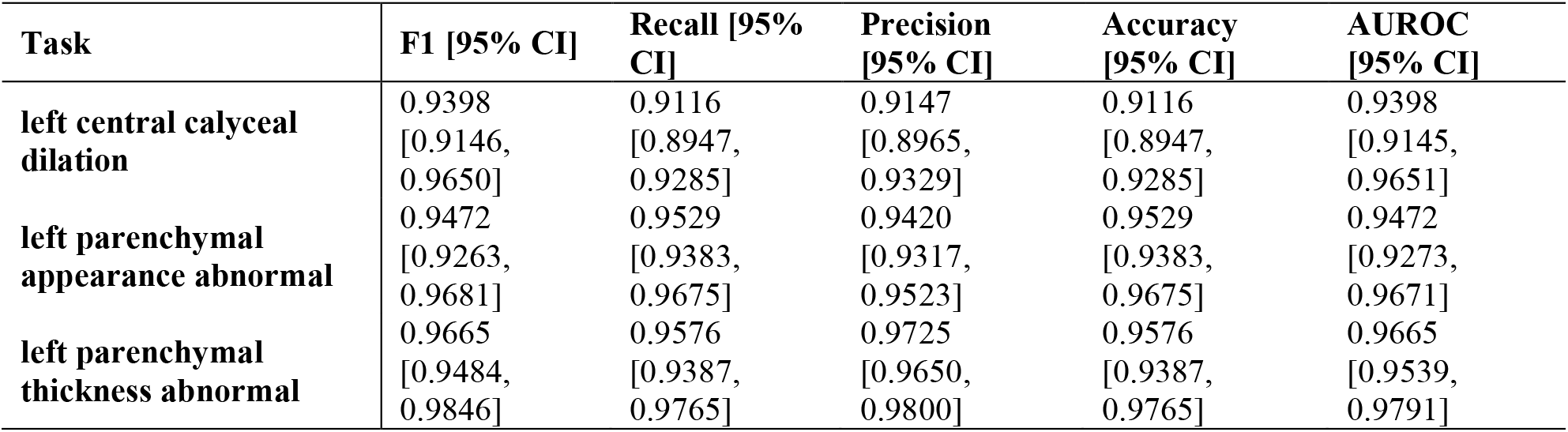
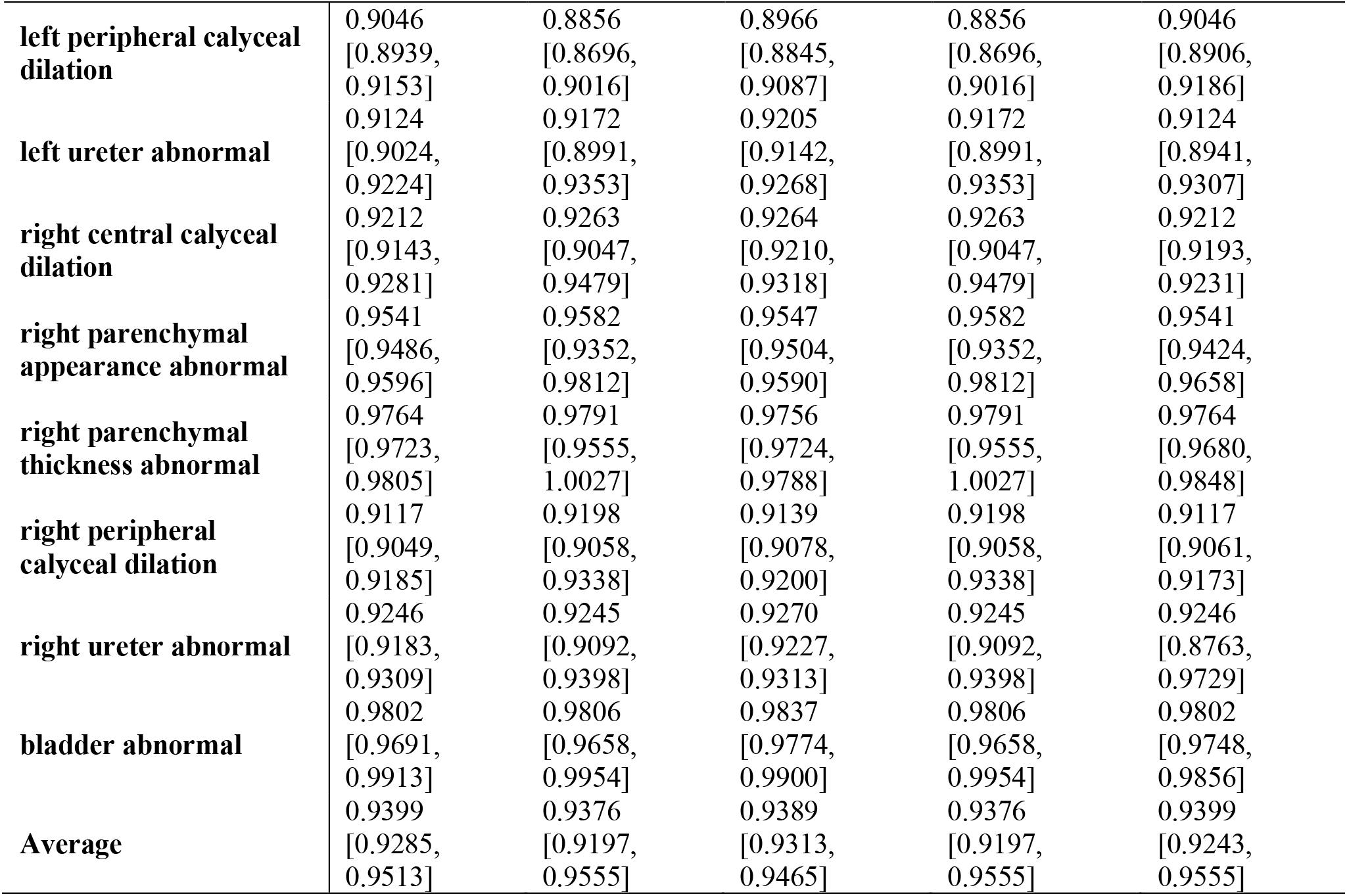
Round 2 model performance.

## REFERENCES

1. Hydronephrosis. In: Tublin M, Borhani AA, Furlan A, Heller MT, eds. Diagnostic Imaging: Genitourinary (Third Edition). Philadelphia: Elsevier; 2016:270–271.

2. Fernbach SK, Maizels M, Conway JJ. Ultrasound grading of hydronephrosis: introduction to the system used by the Society for Fetal Urology. Pediatr Radiol. 1993;23(6):478–480.

3. Nguyen HT, Benson CB, Bromley B, et al. Multidisciplinary consensus on the classification of prenatal and postnatal urinary tract dilation (UTD classification system). J Pediatr Urol. 2014;10(6):982–998.

4. Esteva A, Robicquet A, Ramsundar B, et al. A guide to deep learning in healthcare. Nat Med. 2019;25(1):24–29. doi:10.1038/s41591-018-0316-z

5. Ayala Solares JR, Diletta Raimondi FE, Zhu Y, et al. Deep learning for electronic health records: A comparative review of multiple deep neural architectures. J Biomed Inform. 2020;101:103337. doi:10.1016/j.jbi.2019.103337

6. Liu J, Zhou P, Hua Y, et al. Benchmarking Large Language Models on CMExam --A Comprehensive Chinese Medical Exam Dataset. Published online June 8, 2023. Advances in Neural Information Processing Systems 36. In Press. http://arxiv.org/abs/2306.03030

7. Afzal M, Alam F, Malik KM, Malik GM. Clinical Context–Aware Biomedical Text Summarization Using Deep Neural Network: Model Development and Validation. J Med Internet Res. 2020;22(10):e19810. doi:10.2196/19810

8. Hua Y, Wang L, Nguyen V, et al. A deep learning approach for transgender and gender diverse patient identification in electronic health records. J Biomed Inform. 2023;147:104507. doi:10.1016/j.jbi.2023.104507

9. Singhal K, Azizi S, Tu T, et al. Large language models encode clinical knowledge. Nature. 2023;620(7972):172–180. doi:10.1038/s41586-023-06291-2

10. Devlin J, Chang MW, Lee K, Toutanova K. BERT: Pre-training of Deep Bidirectional Transformers for Language Understanding. ArXiv181004805 Cs. Published online May 24, 2019. Accessed April 19, 2022. http://arxiv.org/abs/1810.04805

11. Alsentzer E, Murphy J, Boag W, et al. Publicly Available Clinical BERT Embeddings. In: Proceedings of the 2nd Clinical Natural Language Processing Workshop. Association for Computational Linguistics; 2019:72–78. doi:10.18653/v1/W19-1909

12. Agrawal M, Hegselmann S, Lang H, Kim Y, Sontag D. Large Language Models are Zero-Shot Clinical Information Extractors. Published online May 25, 2022. doi:10.48550/arXiv.2205.12689

13. Ge Y, Guo Y, Das S, Al-Garadi MA, Sarker A. Few-shot learning for medical text: A review of advances, trends, and opportunities. J Biomed Inform. 2023;144:104458. doi:10.1016/j.jbi.2023.104458

